# Clinical and translational study of ivosidenib plus nivolumab in advanced solid tumors harboring IDH1 mutations

**DOI:** 10.1101/2025.07.19.25331848

**Authors:** Matthew K. Nguyen, Mark Jelinek, Arjun Singh, Brian Isett, Erica S. Myers, Steven J. Mullett, Yvonne Eisele, Jan Beumer, Robert Parise, Julie Urban, Amy Rose, Lorenzo Sellitto, Krishna Singh, Rose Doerfler, Rebekah E. Dadey, Carl Kim, John C. Rhee, Diwakar Davar, Liza C. Villaruz, Melissa Burgess, Jan Drappatz, Megan Mantica, Amy E. Goodman, Hong Wang, Aatur D. Singhi, Jason J. Luke, Dan P. Zandberg, Riyue Bao

## Abstract

**Background:** Cancers that do not respond to immunotherapy typically harbor a non-T cell-inflamed tumor microenvironment (TME), characterized by the absence of type I/II interferon (IFN) signaling and CD8^+^ T cell infiltration. We previously reported *IDH1* somatic mutations were enriched in non-T cell-inflamed tumors across tumor types. Consistent with this, mutant IDH1 (mIDH1) has been demonstrated to drive immune exclusion through metabolic reprogramming of the TME, and IDH inhibition enhanced anti-tumor immunity in preclinical models. Based on these pan-cancer observations, we conducted a Phase II study assessing the preliminary activity of ivosidenib, an IDH1 inhibitor, in combination with nivolumab, an anti-PD1 antibody, in patients with mIDH1 advanced solid tumors (NCT04056910).

**Methods:** Patients with an advanced or refractory solid tumor harboring an *IDH1* mutation, but no prior exposure to IDH1 inhibitor, were enrolled. Participants were administered ivosidenib 500 mg by mouth daily with nivolumab 480 mg intravenously every 4 weeks. Given heterogeneity in tumor types, including some where RECIST response is uncommon (i.e. sarcoma), a composite primary endpoint was utilized including either six-month progression free survival (PFS6) or overall response rate (ORR). Translational analyses included pharmacodynamic, proteomic, and spatial transcriptomic investigations.

**Results:** 15 patients were enrolled (median age, 54 years; female, 53.3%; ECOG 1, 60%; glioma, 46.7%; R132H, 40%). One patient had a partial response (ORR 6.7%) and was without progression at 6 months (PFS6) whereas two other patients achieved PFS6 alone. In total, 3 out of 15 patients met the primary endpoint (3/15; 20%). The median PFS was 1.94 months. The most common treatment adverse events were leukopenia (67%), rash (67%), diarrhea (33%), nausea (27%), and QTC prolongation (27%). Pharmacodynamic studies demonstrated combining ivosidenib and nivolumab significantly decreased the plasma (R)-2HG concentration and correlated with clinical benefit. Serum proteomic and spatial omic analysis suggested immune-modulatory effects of mIDH1 inhibition plus anti-PD1.

**Conclusions:** In treatment refractory mIDH1 solid tumors, the combination of ivosidenib and nivolumab was safe however demonstrated similar anti-tumor activity (predominantly as disease stabilization) compared with that previously described for ivosidenib monotherapy. Translational investigation suggests further evaluation of IDH1 inhibition as a combination partner with immune-checkpoint inhibition may be justified.

**Implications for practice:** This study demonstrates that ivosidenib combined with nivolumab was safe in patients with advanced IDH1-mutant solid tumors. The clinical activity was modest and comparable to ivosidenib monotherapy. Pharmacodynamic and exploratory translational analyses revealed immune-related changes in the tumor microenvironment, suggesting that mutant IDH1 inhibition may modulate immune signaling. These findings support further investigation of IDH1 inhibitors as immunotherapy partners and highlight the importance of integrated translational analyses to inform therapeutic development.

## Background

Immune checkpoint inhibitors (ICI) have transformed cancer treatment. However, patients with a non-T cell-inflamed tumor microenvironment (TME), characterized by the absence of interferon signaling and tumor infiltration by CD8^+^ T-cells, experience limited benefit (1–3). Our group identified that isocitrate dehydrogenase 1 (IDH1) mutations (mIDH1; R132H and R132C) were associated with a non-T cell-inflamed phenotype across cancers (4). mIDH1 confers gain-of-function activity, converting α-ketoglutarate to the oncometabolite (R)-2-hydroxyglutarate [(R)-2HG], which accumulates in tumor cells and subsequently released into the TME (5–7). (R)-2HG inhibits α-ketoglutarate-dependent enzymes, promotes DNA hypermethylation, silences cGAS-STING-IRF3 signaling, and suppresses cytokine/chemokine releases, leading to a reduced CD8^+^ T-cell recruitment and an immunosuppressive TME (8–12). In preclinical tumor models, mIDH1 inhibition reversed these processes, restored anti-tumor immunity, and significantly reduced tumor growth (9).

Based on these mechanistic insights and strong correlation of mIDH1 with the non-T cell-inflamed phenotype across tumor types (**Fig. S1**), we hypothesized that IDH1 inhibition combined with ICI may be an effective treatment strategy for patients with mIDH1 tumors. Here we report a Phase II study evaluating the safety and preliminary anti-tumor activity of the mIDH1 inhibitor ivosidenib plus the anti-PD1 antibody nivolumab in patients with mIDH1 advanced solid tumors, as well as associated pharmacodynamic, serum and tumor based translational correlates.

## Methods

### Patient eligibility

Adult men and women were eligible for enrollment into this trial if they had a histopathologic diagnosis of an advanced solid tumor with documented *IDH1* gene mutation (R132C/L/G/H/S) by sequencing. Patients had to have progressed on appropriate standard of care treatment or for which no curative treatment was available. Patients had to have at least one evaluable and measurable lesion by RECIST v1.1 (solid tumors) or by Response Assessment in Neuro-Oncology (RANO) criteria (glioma). Patients with glioma were required to have diseases that were both WHO 2016 grade ≥ 2 and contrast enhancing. Patients were required to have a good performance status (ECOG PS of 0 or 1) and adequate end organ function. Toxicities associated with prior anticancer therapy must have been resolved to baseline or ≤ grade 1.

Key exclusion criteria included, but were not limited to, having received a prior IDH1 inhibitor; having received systemic anticancer therapy or an investigational agent less than 2 weeks prior to day 1; active autoimmune disease requiring systemic treatment in the past 2 years; or a diagnosis of immunodeficiency. Patients with non-glioma solid tumors must not have received radiotherapy to metastatic sites of disease < 2 weeks prior to day 1 and glioma patients must not have received radiation within 3 months prior. Non-glioma patients must not have undergone hepatic radiation, chemoembolization, or radiofrequency ablation < 4 weeks prior to day 1. Patients with non-glioma solid tumors were ineligible if they had known symptomatic brain metastases requiring steroids. Patients with previously diagnosed brain metastases were eligible for enrollment if they had completed their treatment and had recovered from the acute effects of radiation therapy or surgery prior to study entry, had discontinued corticosteroid treatment for these metastases for at least 1 week, and had radiographically stable disease for at least 1 month prior to being enrolled on the study.

### Clinical trial design

This single-center Phase II clinical trial was designed to assess the preliminary activity of ivosidenib in combination with nivolumab in patients with mIDH1 advanced solid tumors (NCT04056910, **Fig. S2**). Participants were administered ivosidenib 500 mg by mouth daily in combination with nivolumab 480 mg intravenously every 4 weeks. Based on the tumor spectrum of IDH1 mutations in cancer, it was observed that some tumor types are rarely associated with RECIST responses, for example chondrosarcoma. Therefore, a composite primary endpoint was designed to include either of six-month progression free survival (PFS6) or overall response rate (ORR). Utilizing PFS6 as an endpoint was proposed to allow capture of clinical benefit in the form of “cytostatic” effect previously observed and proposed as a priority primary endpoint for soft tissue sarcomas (13, 14). Thus, a participant was considered to meet the primary endpoint if they had PFS6 (scored as either yes or no) or they had at least a partial response (PR; based on RECIST v1.1 in solid tumors or RANO in glioma) after the week 8 scan. This study was conducted in compliance with the Declaration of Helsinki and the International Conference on Harmonization Guidelines for Good Clinical Practice. The University of Pittsburgh institutional review board (IRB) approved this protocol (HCC#19-096). Participants gave informed consent to participate in the study before taking part. All samples have written informed patient consent.

This trial employed an Optimal Simon two-stage design where a probability of a positive outcome < 0.1 would not be promising and a probability of a positive outcome ≥ 0.3 would warrant further interest. In the first stage, 18 participants were proposed to be accrued however the study was eventually discontinued due to lack of clear clinical activity and changes in the landscape of treatment. This included approval of anti-PD1/L1 antibody in the first line of therapy for cholangiocarcinoma as well as the launch of a competing industry sponsored clinical trial of IDH inhibition with dual checkpoint blockade. Trial accrual occurred from March 29^th^, 2021 to October 25^th^, 2023.

### Translational analysis of human specimens from trial

To understand treatment-induced changes in patients, we conducted analyses of longitudinal 2-hydroxyglutarate (2-HG) levels and proteomics in peripheral blood, as well as high-resolution gene expression by spatial transcriptomics in tumor tissues. A full description of the materials and methods is provided in **Supplementary Methods**. In brief:

1. <u>Liquid chromatography tandem mass spectrometry (LC-MS/MS) analysis of 2-HG in</u> <u>human plasma samples.</u> Plasma 2-HG levels were quantified in 33 samples from 10 patients (C1D1, C2D1, C4D1, EOT) using a validated LC-MS/MS assay at the UPMC Hillman Cancer Center Pharmacokinetics and Pharmacodynamics Facility. 4 patients without paired C1D1 and C2D1 samples were not included. After protein precipitation with ice-cold methanol and addition of D₃-2-HG as internal standard (IS), samples were centrifuged, dried, reconstituted, and analyzed using an Agilent 1200 SL HPLC system coupled to a SCIEX 4000 triple quadrupole MS (ESI negative MRM mode: 151.8>133.9, 149.8>132.2, 146.8>129.0 for 2-HG surrogate, D₃-2-HG, and 2-HG, respectively). Chromatography employed an Atlantis dC18 column with acetonitrile/water (0.1% formic acid) gradient. Calibration curves were generated with 2-HG-[¹³C₅] surrogate analyte in blank plasma, using linear regression (1/y² weighting) of surrogate/IS ratios. The assay was linear (30-30,000 ng/mL) and showed high accuracy and reproducibility.
2. <u>Olink proteomic analysis of human serum samples.</u> Olink Target 96 Immuno-Oncology panel (Cat# 95311-A, Olink Proteomics) was used to analyze 14 serum samples (C1D1, C2D1) from 7 patients with cholangiocarcinoma or chondrosarcoma. One microliter of each sample was processed on the Olink Signature Q100 platform, with data normalized to inter-plate controls (IPCnorm) and reported as Normalized Protein Expression (NPX, log_2_ scale). Differential protein abundance between clinical benefit (CB, PFS ≥4m) and no clinical benefit (NCB) groups across timepoints was identified using limma regression (v3.58.1) with interaction design *(∼ Group * Timepoint*) and patient ID as blocking factor. Proteins with nominal *P*<0.05 were selected for exploratory analysis.
3. <u>Visium HD spatial transcriptomics</u>. Pre- and on-treatment FFPE tumor biopsies from one patient with CB were analyzed using Visium HD spatial transcriptomics (10x Genomics, Cat# 1000675). All samples were processed at the same time and assayed on the same Visium HD slide to minimize batch effect. The pre-treatment tumor biopsy was obtained prior to C1D1 and within 28 days of treatment start. The on-treatment tumor biopsy was obtained at C2D1. Other patients did not have biopsies performed or had insufficient tumor tissue, therefore not included on the assay. H&E-guided regions with viable tumor were selected by pathology review. Five-micron sections were mounted on hydrogel-coated slides (Schott Cat# 1800434) and processed per manufacturer protocol (CG000685 Rev B). RNA quality (DV200 ≥30%) was confirmed, and libraries were prepared and sequenced on Illumina NextSeq 2000 (P4 flow cell, ≥275M reads/sample). Reads were aligned to GRCh38-2020-A using SpaceRanger (v3.1.3) and analyzed in Seurat (v5.3.0) at 8μm bin resolution. Bins with nUMI ≥25, nGene ≥20, and log10GenesPerUMI > 0.92 were retained. Normalization used the “LogNormalize” method with scale factor 10,000 in function *NormalizeData*. Samples were integrated via reciprocal PCA, and clustering (30 PCs, resolution=0.5) identified 20 clusters, and visualized by Uniform Manifold Approximation and Projection (UMAP). Marker genes were identified with *FindAllMarkers* (min.pct=0.01, min.cells.feature=3, test.use=wilcox; FDR 0.01), and up to 25 top-ranked markers per cluster were used for biological annotation. Clusters were mapped to H&E images for spatial validation. Tumor and stroma regions were annotated using QuPath (v0.5.1) with pixel classification, and corresponding bins were extracted for compartment-specific gene expression analysis. Differential gene expression (DEGs) between pre- and on-treatment samples were identified using *FindMarkers* (min.pct=0.1, min.cells=200, test.use=wilcox; FDR 0.01, avg_log2FC ≥ log_2_(1.5) or ≤ ─log_2_(1.5)). Pathway enrichment was performed with Enrichr (accessed June 2025) with BioPlanet 2019 curated gene sets. Pseudobulk expression from tumor/stroma compartments was analyzed with xCell (v1.1.0) for digital cytometry, which combines single-sample gene set enrichment analysis (ssGESA) with reference markers and deconvolution approaches to mitigate the issue of marker co-expression on multiple cell types.

### Survival analysis

PFS and OS were estimated with the Kaplan-Meier method using PROC LIFETEST and plotted with PROC SGPLOT in SAS 9.4 (SAS Institute Inc. Cary, NC). Patient 008 had no tumor measurements due to clinical progression prior to follow up scans and was not included in analysis.

### Statistical analysis

For 2-HG data from plasma samples, its abundance was transformed into log_10_ scale and compared between different timepoints of 10 patients by two-sided paired *t*-test. For Olink targeted proteomics (NPX values at log_2_ scale), differentially abundant proteins were identified using limma regression models. For spatial transcriptomics of pre/on-treatment tumors from patient 016, on-*versus* pre-treatment DEGs were detected by Wilcoxon rank-sum test on normalized, unintegrated read counts per Seurat’s best practice. Pathway enrichment was identified using hypergeometric test. All tests are two-sided unless otherwise noted. P-values were adjusted by BH-FDR method when appropriate. Statistical analysis was performed in R (v4.4.2).

## Results

### Baseline Patient Characteristics

A total of 15 patients were enrolled, with baseline characteristics described in **Table 1**.

**Table 1.** Baseline demographics of patients.

|  | Number of Participants, n = 15 |
| --- | --- |
| **Age at Enrollment (Median (IQR))** | 54 [50,60] |
| **Sex (%)** |  |
| Female | 8 (53.3) |
| Male | 7 (46.7) |
| **Race (%)** |  |
| Black or African American | 1 ( 6.7) |
| White | 14 (93.3) |
| **Ethnicity (%)** |  |
| Hispanic or Latino | 1 ( 6.7) |
| Non-Hispanic | 14 (93.3) |
| **ECOG Performance Status (%)** |  |
| 0 | 6 (40.0) |
| 1 | 9 (60.0) |
| **Disease Histology (%)** |  |
| CHOLANGIOCARCINOMA | 4 (26.7) |
| CHONDROSARCOMA | 3 (20.0) |
| GLIOMA | 7 (46.7) |
| COLORECTAL ADENOCARCINOMA | 1 ( 6.7) |
| **IDH1 Mutation (%)** |  |
| R132C | 5 (33.3) |
| R132H | 6 (40) |
| R132G | 4 (27.7) |
| **Prior Systemic Therapy<sup>1</sup> (%)** | 13 (86.7) |
| Median No. of lines (range) | 2 (0 - 3) |
| Median No. of lines (range) - Cholangiocarcinoma | 1.5 (1 - 3) |
| Median No. of lines (range) - Chondrosarcoma | 0 (0 - 3) |
| Median No. of lines (range) - Glioma | 2 (1 - 4) |
| Median No. of lines (range) - Colorectal Adenocarcinoma | 2 (0) |
| **Prior Surgery** | 11 (73.3) |
| **Prior Radiotherapy** | 9 (60) |

[1] No patients received checkpoint inhibition or an IDH1 inhibitor prior to enrollment on the study.

The median age was 54 years (range: 50 - 60) with 46.7% male and 53.3% female, predominantly of white race (93.3%). 60% of patients had an ECOG performance status of 1 (capable of performing most daily activities with some limitations). Tumor histologies were glioma (46.7%), cholangiocarcinoma (26.7%), chondrosarcoma (20.0%) and colorectal adenocarcinoma (6.7%). mIDH1 included R132H (40%), R132C (33.3%), and R132G (27.7%). Median lines of systemic therapy amongst all disease histologies were 2, and most patients received prior surgery or radiotherapy. All gliomas included in this study were high-grade (at least grade 3), contrast enhancing, and had undergone a median of 2 lines of systemic therapy.

### Treatment-Related Adverse Events

Adverse events (AE) are described in **Tables S1** and **S2**. All patients enrolled experienced an adverse event of any grade, with 13 (87%) experiencing a treatment-related adverse event (TRAE) of any grade. Of these TRAEs, 27% were grade 3 or higher. No dose-limiting toxicities (DLT) were observed and there were no fatal TRAEs. The most common TRAEs were leukopenia and rash. The most common grade 3 or higher TRAEs were leukopenia, QTC prolongation, anemia, and hyperthyroidism (**Table 2; Table S3**).

**Table 2.**
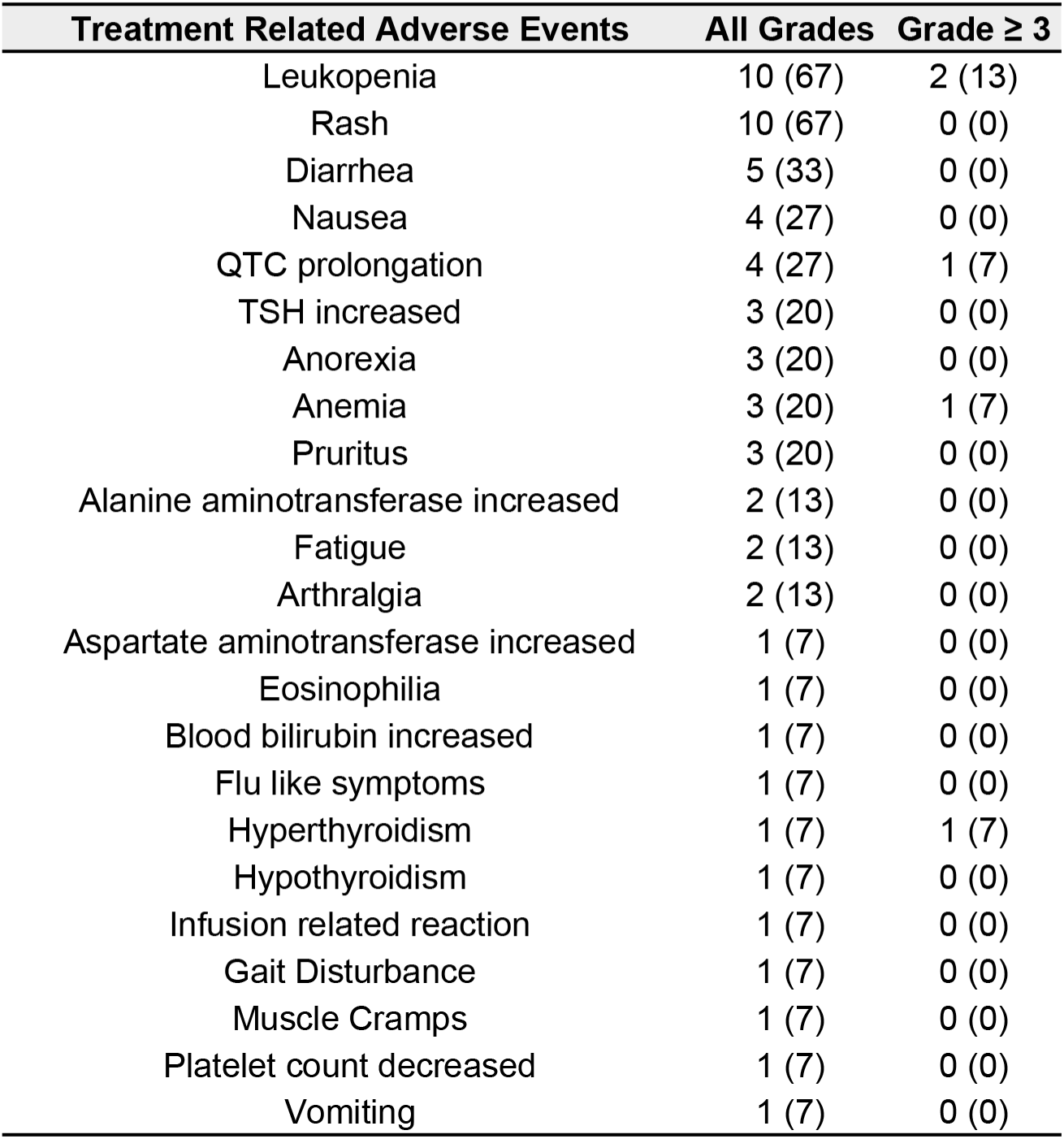
Treatment-related adverse events. Overall adverse events and treatment-related adverse events are summarized with frequencies and percentages in parentheses.

### Treatment Outcome

Out of 15 patients, 14 had tumor measurements with an ORR of 6.67%. One patient had no responses recorded due to clinical progression prior to any follow up imaging. We observed 1 PR (6.67%), 6 stable disease (SD; 40.00%), and 8 progressive disease (PD; 53.33%) 8 weeks after initiation of treatment. One patient with chondrosarcoma (013) had PR. All four patients with intrahepatic cholangiocarcinoma had SD (**Fig. 1A-B**). Three patients met the composite primary endpoint of ORR or PFS6 (3/15; 20.00%). The median PFS was 1.94 months (95% CI 1.61 – 3.68) and median OS was 10.26 months (95% CI 5.10 – 19.66) (**Fig. 1C-D**).

**Figure 1.**
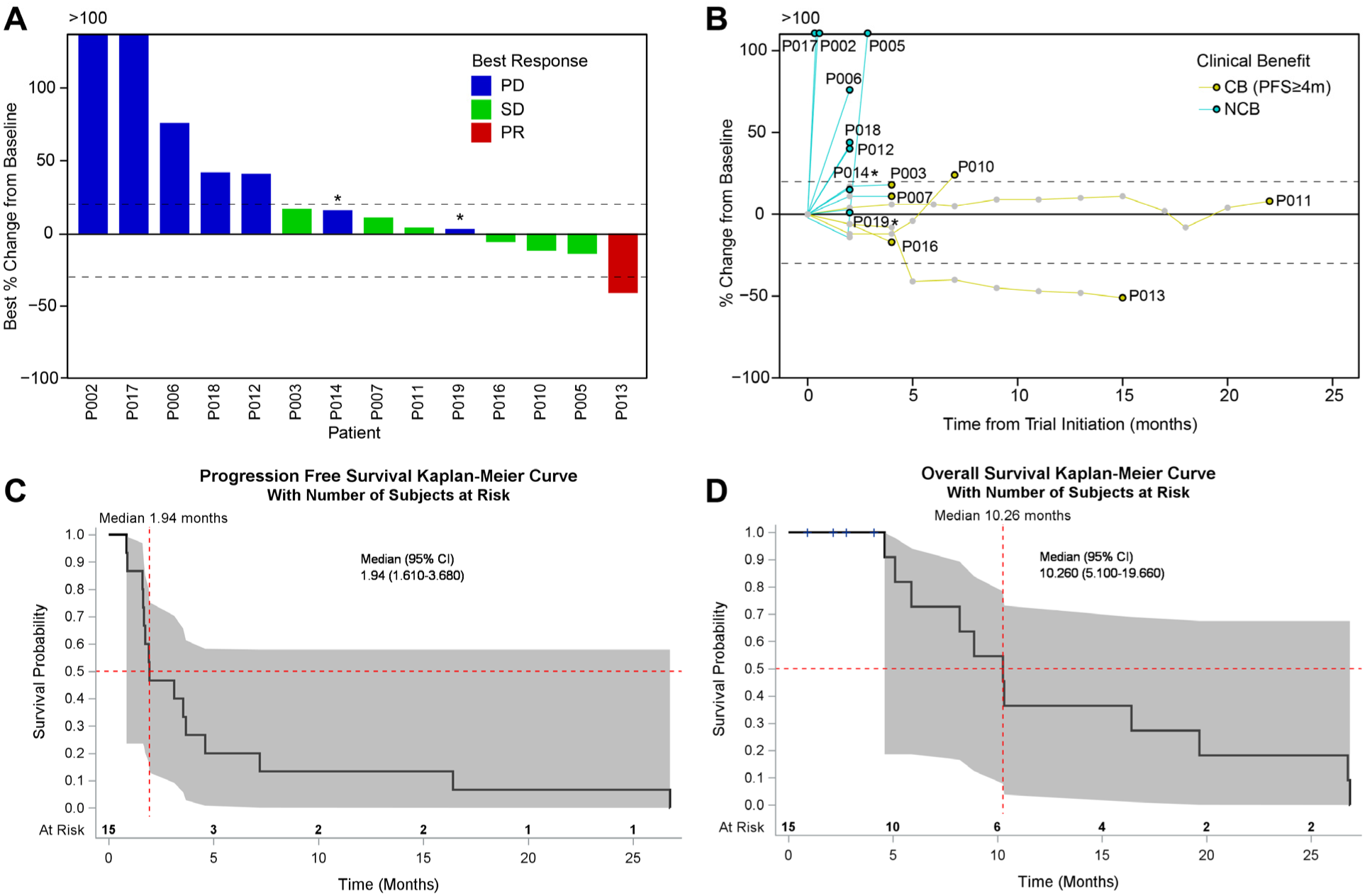
Clinical outcome of ivosidenib plus nivolumab in 14 patients with mIDH1 advanced solid tumors. (**A**) waterfall plot showing best response and best percentage of tumor size change from baseline. Color indicates best response groups. Dotted lines denote cutoffs for PD and PR at +20 and -30 percent. * Asterisk denotes: two patient labeled as PD, one had progression of NT5 lesion and withdrew consent at that time; the other had clinical progression. Both came off treatment. (**B**) spider plot showing percentage of tumor size change from baseline over time. Colored lines indicate clinical benefiters (CB, PFS≥4m), and non-clinical benefiters (NCB). n=14 in **A** and **B**. One patient had no responses recorded due to clinical progression prior to any follow up imaging and not shown (**C-D**). Kaplan-Meier estimates of (**C**) Progression-free survival and (**D**) Overall survival. Red dotted lines indicate the median and shaded indicates the 95% Hall-Wellner confidence band.

Despite the primary endpoint including PFS6, we were interested in detailing the outcomes of any patients who might be considered to derive clinical benefit. Therefore, we specifically call out those with at least PFS ≥ 4 months as possible CB (n=4, 1 chondrosarcoma and 3 cholangiocarcinoma; detailed clinical characteristics described in **Table 3**). All four patients harbored either an R132C or R132G IDH1 mutation.

**Table 3.** Characteristics of patients who experienced clinical benefit.

| Patient ID | Histology | Lines of Systemic Therapy | IDH1 Mutation | Other Biomarkers | PFS/time on treatment (months) | Best Overall Response | Extent of Disease Prior to Enrollment |
| --- | --- | --- | --- | --- | --- | --- | --- |
| 011 | Intrahepatic Cholangiocarcinoma | 1 | R132C | BRAF K601E, ARID1A R1202Q, TMB 3.83 mut/Mb, MSS | 18 | SD | Liver |
| 016 | Intrahepatic Cholangiocarcinoma | 3 | R132C | TP53 H179Q, ERCC2 L461V, KIT D496N, 15.7 Muts/Mb, MSS | 4 | SD | Liver, gastro-hepatic lymph node, peri-portal and porto-caval lymph node, retro-caval and aorta-caval lymph nodes, right cardio-phrenic lymph node, peritoneal fluid, pericardial fluid |
| 010 | Intrahepatic Cholangiocarcinoma | 1 | R132G | KRAS G12D, ATRX G1219A, CREBBP M866I, 8.9 Muts/Mb, MSS | 7 | SD | Liver, Porta-caval lymph node, retroperitoneal lymph nodes, lung, cardio-phrenic lymph nodes. |
| 013 | Chondrosarcoma | 0 | R132G | N/A (Only IDH mutations tested) | 12 | PR | Right scapula, lung, left adrenal, spleen, right supraclavicular lymph nodes |

The PR observed during this trial was the only patient with chondrosarcoma (013) that clinically benefitted. This was a patient with conventional chondrosarcoma of the right scapula. Pathology demonstrates a grade 3 chondrosarcoma with IDH1 R132G mutation identified. In contrast, the other two patients with chondrosarcoma had dedifferentiated histology. This patient enrolled on the trial after recurrence in the bilateral lungs and right proximal upper extremity/shoulder. The patient received 12 cycles of treatment on protocol. Ultimately, the patient was discontinued from the trial due to intolerable grade 2 arthralgias/arthritis. Restaging scans 2 months after the end of trial demonstrated ongoing partial response with a 51% decrease of the target lesions compared to baseline.

### Ivosidenib plus Nivolumab suppresses (R)-2-hydroxyglutarate in plasma

Prior pharmacodynamic studies have shown that plasma (R)-2HG levels were inhibited by as much as 98% compared to baseline after one week of continuous ivosidenib dosing and persisted at C2D1 (5). To confirm that ivosidenib and nivolumab decreased (R)-2HG concentration levels, we collected plasma specimens at C1D1, C2D1, C4D1 as well as EOT (**Fig. S2**). In 10 patients with paired C1D1 and C2D1 plasma (3 chondrosarcoma, 4 cholangiocarcinoma, and 3 glioma), (R)-2HG was significantly reduced at C2D1 relative to C1D1 (*P*=0.0077) (**Fig. 2A**). In 7 patients with cholangiocarcinoma or chondrosarcoma, this pattern was observed in CB (n=3; *P*=0.035) but not in NCB (n=4; *P*=0.085) (**Fig. 2B**). A higher reduction in C2D1-C1D1 (R)-2HG levels was detected in CB compared to NCB (69.70%±8.45 and 60.56%±11.8, respectively; mean±S.E.M.). At the time of progression or EOT, a trend toward increasing concentrations of (R)-2HG back to baseline was observed (**Fig. 2B**). Our study shows that the combination of ivosidenib and nivolumab significantly reduced (R)-2HG levels in patients, with an even greater reduction observed in patients who experienced a clinical benefit. However, this reduction was not as pronounced as the near-98% inhibition of (R)-2HG previously reported when ivosidenib was used alone in earlier studies (5).

**Figure 2.**
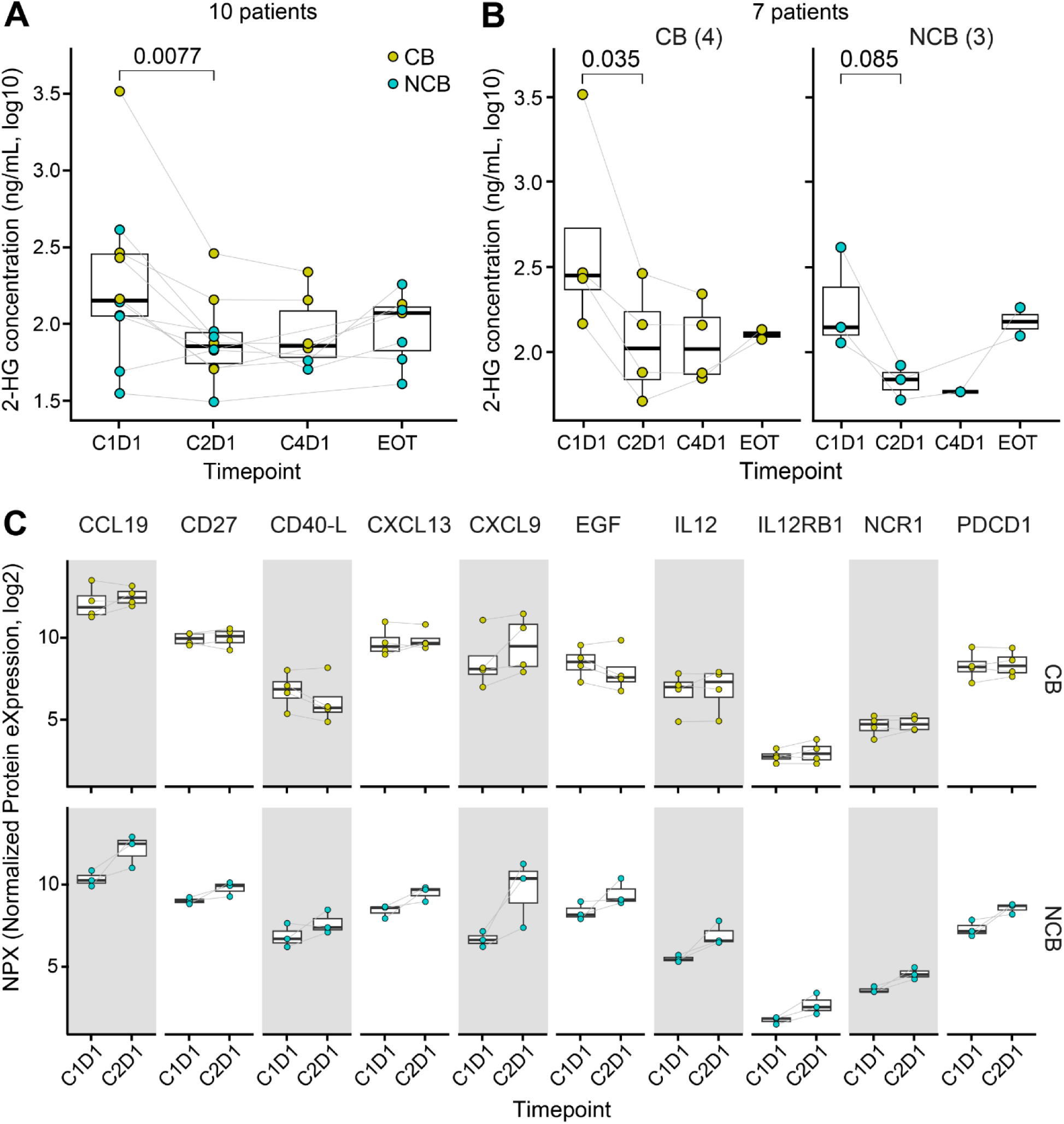
Peripheral blood marker changes upon Ivosidenib plus nivolumab. (**A**) Plasma (R)-2HG levels. N=10 patients shown. (**B**) Plasma (R)-2HG concentration stratified by clinical benefiters *versus* non-benefiters. N=7 patients shown. (**C**) Serum protein expression in clinical benefiters and non-clinical benefiters. Ten proteins at nominal *P*<0.05 are shown. Two-sided Welch’s two-sample *t*-test was used in **A** on log_10_ transformed data. Limma regression models were used in **B**. C1D1 = cycle 1 day 1. C2D1 = cycle 2 day 1. C4D1 = cycle 4 day 1. EOT = end of treatment. Color denotes clinical benefiters (CB, PFS≥4m) and non-clinical benefiters (NCB).

### Proteomic analysis identifies pro-inflammatory protein expression with mIDH1 inhibition plus ICI

To determine longitudinal proteomic changes in peripheral blood, C1D1 and C2D1 serum samples from 7 patients with cholangiocarcinoma (n=4) or chondrosarcoma (n=3) were analyzed using Olink 96-plex targeted immuno-biology panel. We identified ten proteins with treatment-induced abundance differences between CB and NCB, including CCL19, CD27, CD40L, CXCL13, CXCL9, EGF, IL-12, IL-12RB1, NCR1, and PDCD1 (nominal *P*<0.05; **Fig. 2C**). Contrary to our initial hypothesis that these pro-inflammatory markers would increase in patients with clinical benefit, these proteins were consistently upregulated in C2D1 relative to C1D1 specifically in NCB. Conversely, CB showed no distinct trend of increase in these same proteins. Among these, CXCL9 showed the largest increases in protein expression in both groups, albeit with different magnitudes or patterns between CB and NCB (average log_2_(fold change)=1.01 and 3.00, respectively; **Fig. 2C**).

### Spatial transcriptomics reveal immune gene expression changes upon treatment

To investigate treatment-induced TME changes, we analyzed paired pre- and on-treatment specimens from one patient (016) who experienced CB and had sufficient tumor tissues for Visium HD spatial transcriptomics. This patient had intrahepatic cholangiocarcinoma, a R132C IDH1 mutation, and an overall best response of SD. Due to low gene detection at the original 2μm bin size, we aggregated data into 8μm bins to improve expression capture. UMAP-based clustering revealed 20 spatially distinct neighborhoods (C00-C19; **Fig. 3A**), with 12 clusters retained for biological interpretation describing malignant, immune, endothelial, or fibroblast activity (**Fig. S3**). Both samples showed similar cluster distribution by UMAP (**Fig. 3A, left**) and bin proportions (**Fig. S4A-B**), suggesting minimal batch effect. Tumor cell-rich neighborhoods were predominantly in clusters 00 and 04, with others as stromal neighborhoods (**Figs. S05 and S06**). Of interest, C00 were enriched at tumor-stroma boundaries (**Fig. 3B, middle, orange**) and C04 in tumor core (**Fig. 3B, middle, blue**).

**Figure 3.**
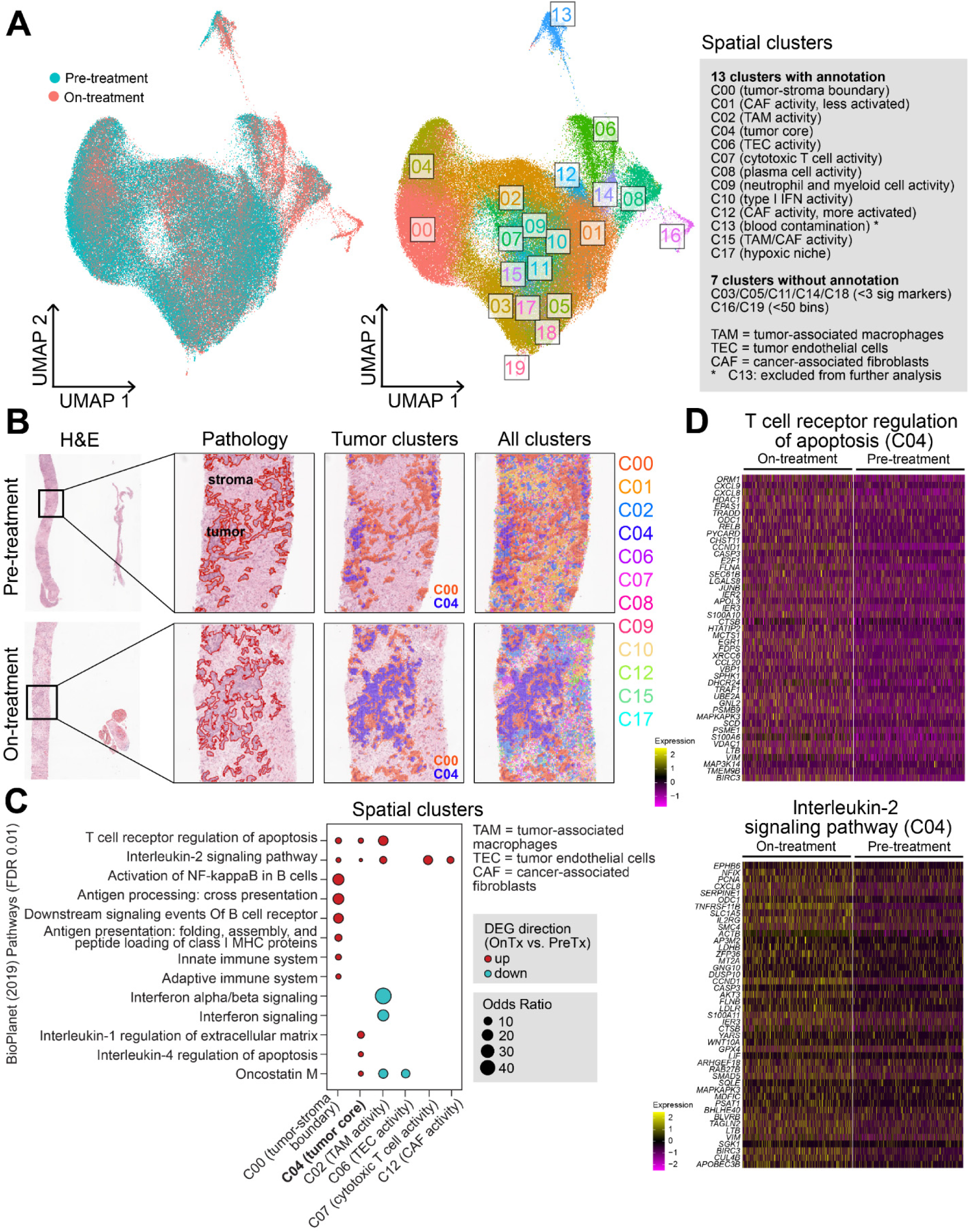
TME changes by spatial transcriptomics upon Ivosidenib plus nivolumab. (**A**) Clusters by UMAP showing distinct cellular neighborhoods with biological interpretation. (**B**) Representative examples of pathology tumor/stroma annotation and spatial clusters from pre- and on-treatment samples. (**C**) Immune-related pathway enrichment in each spatial cluster (FDR-adjusted *P*<0.01). Clusters without significant pathways are not shown. (**D**) Gene expression heatmap of two immune pathways from **C**, showing that in tumor core (C04), on-treatment sample has significantly higher pathway gene expression compared to pre-treatment sample (FDR-adjusted *P*<0.01). Two-sided hypergeometric test was used in **C**, two-sided Wilcoxon rank-sum test was used in **D**. BH-FDR adjustment of p-values was used in multiple comparisons.

We and others have shown that IDH activation is associated with immune exclusion (4), and that IDH inhibition enhances anti-tumor immunity in preclinical models (9). To investigate the immune impact of IDH inhibition plus anti-PD1 in human tissue, we identified DEGs between on- and pre-treatment tissues within each cluster (FDR-adjusted *P*<0.01), and performed pathway analysis with curated gene sets (BioPlanet 2019). Immune-related pathways, including T-cell receptor regulation of apoptosis and IL-2 signaling, were upregulated in both tumor clusters (C00, C04) as well as in cytotoxic T cell (C07) and cancer-associated fibroblasts (CAF; C12) neighborhoods (**Fig. 3C-D**). In contrast, tumor-associated macrophage (TAM)-rich neighborhoods (C02) showed downregulation of IFN-α/β signaling genes (e.g., *IFITM3, OAS1-3, MX1, ISG15*), suggesting potential immunosuppression.

Using an orthogonal approach by xCell on pseudobulked gene expression from tumor and stromal compartments, we found higher enrichment scores for T cell and dendritic cell (DC) subsets in the on-treatment sample, alongside an increase in M2-like macrophages (**Fig. S7**). These findings indicate that ivosidenib plus nivolumab enhances anti-tumor immunity, but this may be offset by increased immunosuppressive macrophages, potentially contributing to the SD rather than PR observed in this patient.

## Discussion

Here we report a phase II clinical trial investigating the combination of ivosidenib, an IDH1 inhibitor, with the anti-PD1 antibody nivolumab in patients with IDH1 mutant advanced solid tumors. We observed a safety profile consistent with the known toxicity spectrum of the individual agents but no clear suggestion of combinatorial benefit. Three of 15 patients met the composite primary endpoint of ORR or PFS6, with a single PR.

We and others have observed pan-cancer associations between *IDH1* mutations, the non-T cell-inflamed TME and immunotherapy resistance. mIDH1 leads to production of the oncometabolite (R)-2HG which alters DNA methylation and suppresses interferon signaling (11, 12). (R)-2HG in the TME may impair production of IFNγ and IL-2 by CD4^+^ and CD8^+^ T cells as well as impacting myeloid cell populations, for example immunosuppressive glioma-associated macrophages generated due to altered tryptophan metabolism (15, 16). In human glioma, an inverse relationship between (R)-2HG concentrations and expression of interferon and antigen presentation signaling pathways, as well as CD3^+^ and CD8^+^ T-cells, has been observed (17). In cholangiocarcinoma, inhibition of mIDH1 correlates with induction of interferon responsive molecules, such as PD-L1, PD1, and VISTA/BY-H5, on tumor infiltrating immune cells and lower lymphocytes counts within mIDH1 versus wildtype tumors (19). Overall, these studies emphasize that mIDH1 likely has an immunomodulatory role and inhibition may therefore have potential as a combinatorial partner for ICI. Despite this rationale, the clinical outcomes from our study are more aligned with those seen in clinical trials evaluating the efficacy of ivosidenib monotherapy. The ClarIDHy trial assessed the use of ivosidenib in patients with advanced mIDH1 cholangiocarcinoma and reported a median PFS of 2.7 months and ORR of 2% (20). In gliomas, ivosidenib demonstrated a median PFS of 1.4 months (95% CI, 1.0 to 1.9 months) in the enhancing by MRI cohort vs. 13.6 months (95% CI, 9.2 to 33.2 months) in the non-enhancing cohort (21). The gliomas treated in our study were predominately enhancing and had undergone more lines of systemic therapy as compared to the approved setting where vorasidenib is used for Grade 2 astrocytoma or oligodendroglioma (22).

Noting the heavily pre-treated nature of our cohort, we were interested in further investigating the pharmacodynamic and translational impact of mIDH1 inhibition with anti-PD1. Consistent with prior studies, we observed significant suppression of (R)-2HG from C1D1 to C2D1 and that clinical benefit was associated with greater degrees of (R)-2HG suppression after the first cycle. In contrast with prior studies, the degree of (R)-2HG suppression observed was substantially lower than that previously reported (5). The reason for this is unclear and may contribute to the minimal clinical efficacy observed. Despite this lower (R)-2HG suppression, serum proteomic analyses did reveal upregulation of ten proteins involved in immune signaling and regulation, however this was not associated with clinical benefit. Further studies correlating these proteomic changes with TME analyses and more extensive longitudinal sampling would be warranted surrounding the combination of mIDH1 inhibition plus ICI.

Despite an initial plan for pre- and on-treatment tumor sample collection, the ability to actualize this was limited. Spatial omic characterization of the TME prior to and during therapy for one patient with cholangiocarcinoma, who had clinical benefit, was obtained. Analysis of the TME demonstrated increased gene expression of immune-related pathways, such as T cell receptor regulation of apoptosis and IL-2 signaling, in multiple spatially distinct tumor and stroma clusters. Meanwhile, immunosuppressive signals including M2-like macrophages were observed. Collectively, these data suggest that the combination of mIDH1 inhibition and anti-PD1 may result in revitalization of the T-cell inflamed TME, though further work is needed to validate this observation.

Limitations of this study may include but not be limited to the single arm design and small sample size. Based on the observation for correlation of mIDH1 and the non-T-cell-inflamed TME across tumor types, an underlying hypothesis for the study was that mIDH1 may be an immunotherapy combination target independent of cancer histology. In retrospect, including any tumor harboring IDH1 mutation may have hindered the study, especially due to the inclusion of gliomas (47% of the study population) where ivosidenib has minimal activity in enhancing disease. Due to limited biopsy tissue available, we were unable to perform extensive immunohistochemistry (IHC) validation on immune cell findings from the Visium HD studies. With this limitation, we analyzed the immune impact of IDH1 inhibition with anti-PD1 via two computational methods including DEG pathway enrichment analysis and digital cytometry (xCell). Given our limited sample size, the clinical and translational data produced from this study are exploratory but may be hypothesis generating for the field.

In conclusion, this phase II trial of ivosidenib with nivolumab demonstrated an expected toxicity spectrum but no obvious differentiation from ivosidenib monotherapy. Exploratory translational analysis suggests that ivosidenib plus nivolumab may have immuno-modulatory impact however further studies, will be required to better assess the utility of this combination. A clinical trial evaluating the efficacy of mutant IDH1 inhibition with dual checkpoint blockade in cholangiocarcioma is underway (23).

## Data Availability

All data relevant to the study are included in the article or uploaded as supplementary information. Other data will be provided upon request from the corresponding author.

https://github.com/HCC-data-sciences-pub/IDH1iplusnivo-data-analysis

## Author Contributions

Conception and design: R.B., J.J.L.; Development of experiments: R.B.; Performance of experiments: E.S.M., S.J.M., Y.E., J.B., R.P., K.S., R.E.D., B.D., C.K.; Collection and assembly of data: R.B., J.J.L., M.K.N., M.J., L.S.; Data analysis and interpretation: R.B., J.J.L., D.P.Z., M.K.N., M.J., A.S., B.I., E.S.M., S.J.M., Y.E., J.B., H.W., A.D.S.; Consent, actual/referral of patients and trial administration: J.J.L., J.U., A.R., L.S., J.C.R., D.D., L.C.V., M.B., J.D., M.M., A.E.G.; Manuscript writing: R.B., J.J.L., M.K.N., A.S., M.J.; Manuscript editing: R.B., J.J.L., D.P.Z.; All authors reviewed and approved the manuscript.

## Acknowledgment

We thank the patients and families for their participation in this study. We thank Dr. Fangping Mu for technical assistance at The University of Pittsburgh Center for Research Computing (CRC) high-performance computing clusters (HPC). This project used the UPMC Hillman Cancer Center Cancer Bioinformatics Facility (CBS), Biostatistics Facility (BC), Translational Oncologic Pathology Services (TOPS), and Cancer Pharmacokinetics and Pharmacodynamics Facility (CPPF). Histology sectioning was performed by the Rangos Histology Core Facility, a shared facility in the Rangos Research Center at the University of Pittsburgh. 10x Visium library generation and Illumina sequencing were performed by the Health Sciences Sequencing Core (RRID:SCR_023116) at UPMC Children’s Hospital of Pittsburgh, Rangos Research Center. Olink assay was performed by Health Sciences Mass Spectrometry Core (RRID:SCR_025222). Services and instruments used in this project were graciously supported, in part, by the University of Pittsburgh, the Office of the Senior Vice Chancellor for Health Sciences, the Department of Pediatrics, the Institute for Precision Medicine, and the Richard K Mellon Foundation for Pediatric Research. **Funding information.** This work was supported by National Institutes of Health (NIH) grant UM1CA186690 (J.J.L., J.H.B.), and in part by National Cancer Institute (NCI) through the UPMC Hillman Cancer Center CCSG award P30CA047904 (R.B.), NCI grant R50CA211241 (J.H.B.), NIH grants S10OD0234402 and S10OD032141 (PI Gelhaus for both), and The University of Pittsburgh CRC through the resources provided specifically the HTC clusters supported by NIH S10OD028483. Bristol Myers Squibb and Servier provided drug substance and funding for the clinical trial but had no input on the study design or the interpretation of the results.

