## Supplementary Methods and Figures 1-7 for "Clinical and translational study of ivosidenib plus nivolumab in advanced solid tumors harboring IDH1 mutations"

This document contains the full description of **Methods** for the manuscript, **Supplementary Figures 1 to 7** and **Title of Supplementary Tables**. The **Supplementary Tables 1 to 3** are supplied as separate spreadsheets**.**

**Supplementary Methods**

**Liquid chromatography tandem mass spectrometry (LC-MS/MS) analysis of 2-Hydroxyglutarate in human plasma samples**

An LC-MS/MS assay was performed at the UPMC Hillman Cancer Center Cancer Pharmacokinetics and Pharmacodynamics Facility to quantify 2-hydroxyglutarate (2-HG) in 33 plasma samples (10 C1D1, 10 C2D1, 6 C4D1, 7 EOT) from 10 patients, including 7 patients with cholangiocarcinoma (n=4) or chondrosarcoma (n=3). Four patients without paired C1D1 and C2D1 samples were not included in the assay. The analytical platform consisted of an Agilent (Palo Alto, CA) 1200 SL series thermostatted autosampler, binary pump system, and column heater, coupled to a SCIEX 4000 triple quadrupole mass spectrometer (SCIEX, Toronto, Canada) operated in ESI negative MRM mode. The mass transitions monitored were 151.8>133.9, 149.8>132.2 and 146.8>129.0 for 2-HG surrogate, $D_3$-2-HG and 2-HG, respectively. Chromatographic separation was achieved using an Atlantis dC-18, 3μm, 2.1 x 50 mm (Waters Corporation, Milford, MA) column with a gradient consisting of HPLC grade (Fisher Scientific, Fairlawn, NJ) acetonitrile and water both with 0.1% formic acid. For sample preparation, 50 μL of human plasma was aliquoted into a microcentrifuge tube. Ten microliters of deuterated 2-hydroxyglutarate ($D_3$-2-HG) was added as the internal standard (IS), followed by 250 μL of ice-cold methanol to precipitate plasma proteins. After vortexing, samples were centrifuged at 17,200 × *g* for 10 minutes at 4°C. The resulting supernatant was transferred to a clean borosilicate 12 x 75 mm glass tube and evaporated to dryness under a stream of nitrogen. The dried extract was reconstituted in 100 μL of HPLC grade water, and a 5 μL aliquot was injected into the LC-MS/MS system. Since 2-HG is an endogenous compound, 2-Hydroxyglutarate-[$^{13}C_5$] disodium salt was used as a surrogate analyte for calibration curve generation. Calibration standards were prepared in blank plasma using the surrogate, and the ratio of the surrogate to the internal standard response was plotted against nominal concentrations. A linear regression model, not forced through the origin and weighted by $1/y^2$, was used to fit the calibration curve. The slope and intercept of the calibration curve were recorded for each analytical run, and the same parameters were used to back-calculate 2-HG concentrations in study samples using the measured 2-HG/IS response ratio. The assay was linear from 30-30,000 ng/mL and provided robust, accurate, and reproducible quantitation of plasma 2-HG concentrations.

**Olink proteomic analysis of human serum samples**

Olink target 96 immuno-oncology proximity extension assay (Olink Proteomics, Boston, MA, part of ThermoFisher Scientific, Cat# 95311-A) was performed in 14 serum samples (7 C1D1, 7 C2D1) from 7 patients with cholangiocarcinoma (n=4) or chondrosarcoma (n=3). One microliter of each serum sample was assayed on an Olink Signature Q100 instrument (Olink Proteomics) to quantify the protein abundance. The resulting data were collected using the manufacturer's software (Olink Signature software 2.0) and the intensity was normalized to inter-plate controls (IPCnorm). Results were reported as Normalized Protein Expression (NPX), an arbitrary unit on the log2 scale representing protein abundance.

Comparing patient groups with clinical benefit (CB, PFS≥4m) to those without clinical benefit (NCB), treatment-induced (C2D1 to C1D1,*Timepoint*) protein abundance changes in NPX were identified using limma[1] regression models (v3.58.1), with design matrix function as "model.matrix(~ *Group* * *Timepoint*)" and "*Patient ID*" as blocking factor. Proteins of interest were filtered by nominal $P<0.05$ for exploratory analysis.

**Visium HD library preparation and sequencing of tumor FFPE tissues**

Spatial transcriptomics by Visium HD was performed on pre/on-treatment tumor biopsies from 1 patient (016), who experienced CB and sufficient viable tumor tissues were identified by a pathologist (A.D.S.). The pre-treatment tumor biopsy was obtained prior to C1D1 and within 28 days of treatment start. The on-treatment tumor biopsy was obtained at C2D1. Other patients did not have biopsies performed or had insufficient tumor tissue, therefore not included in the assay. All samples were processed at the same time and assayed on the same Visium HD slide to minimize batch effect. Samples were assessed for RNA quality, and all FFPE samples had a DV200 score above the minimum threshold ≥30%. Spatial transcriptomic libraries were generated with Visium HD Human Transcriptome gene expression kit (10x Genomics: 1000675). 5 micron FFPE sections were placed on Schott Nexterion H-3D Hydrogel Coated Slides (Schott North America: 1800434) to minimize the risk of tissue detachment and processed through the 10x Visium HD for FFPE protocol according to the manufacturer's instructions (CG000685 Rev B). H&E images were taken at 40x magnification using a Leica Aperio CS2 slide scanner (Leica Biosystems). Library QC was completed with an Agilent TapeStation 4150. Libraries were normalized and pooled to 2nM prior to loading on an Illumina

NextSeq 2000, using a P4 100 flow cell with a target of a minimum of 275 million reads per sample for transcriptomic libraries.

**Visium HD data analysis**

Visium HD reads were aligned to human transcriptome GRCh38-2020-A with 10x Genomics Space Ranger (v3.1.3), and analyzed using Seurat[2] (v5.3.0). Data QC, filter, and normalization: Both samples were merged into one object and analyzed based on 8μm resolution after aggregation from 2μm bins. Data were filtered to keep bins with nUMI ≥ 25, nGene ≥ 20, log10GenesPerUMI > 0.92, and only genes expressed in at least one bin. Feature count matrix was normalized using "LogNormalize" method with "scale.factor=10000" in function *NormalizeData*, which divides counts for each cell by the total counts for that cell and multiplied by the scale.factor, followed by natural-log transformation using log1p. Clustering and annotation: Samples were integrated using anchor-based reciprocal PCA (RPCA) as implemented in Seurat on 2000 variable features. The integrated object was used to scale data, perform PCA, bin clustering (30 PCs at 0.5 resolution), and generate Uniform Manifold Approximation and Projection (UMAP) for data visualization. 20 clusters were generated, and representative markers of each cluster were detected using function *FindAllMarkers*, with parameters "min.pct = 0.01, min.cells.feature = 3, test.use = wilcox" and other settings as default, and filtered at FDR-adjusted $P$<0.01. Biological annotation of each cluster was conducted using up to 25 top significant markers within each cluster (ranked by p-value from smaller to bigger), with manual curation using violin plots and expression heatmaps across clusters and further inspected by mapping annotated clusters to the H&E slides. Annotation labels were added to 13 out of 20 clusters; 5 clusters of 0-2 significant markers (C03/C05/C11/C14/C18) were considered undefined; 2 clusters (C16/C19) of less than 50 bins were excluded. C13 (blood contamination) was excluded from further analysis. Pathology annotation: On the 40x HE images, tumor and stroma compartments were created within QuPath (v0.5.1) by first manually creating a training set within each image and then extending the prediction across the entire image by pixel classification. Regions smaller than 1000 pixels were merged. The resulting tumor and stroma masks were used to extract the 8 μm bins within each compartment and create a filtered Seurat object for subsequent analysis. DEG detection: Within each cluster, DEGs were identified using normalized, unintegrated expression in on- relative to pre-treatment samples using function *FindMarkers*, with parameters "min.pct = 0.1, min.cells.feature = 200, test.use = wilcox" and other settings as default, and filtered at FDR-adjusted $P$<0.01, and avg_log2FC ≥ $\log_2(1.5)$ or ≤ $-\log_2(1.5)$. Pathway enrichment of significant

DEGs of each cluster was detected using Enrichr[3] (accessed June 2025) with BioPlanet 2019[4] gene sets. Digital cytometry: Pseudobulk gene expression of tumor and stroma compartments was created using function *AggregateExpression*, and used to identify the enrichment of defined cell types in each compartment using xCell[5] (v1.1.0). xCell combines single-sample gene set enrichment analysis (ssGSEA) with reference markers and deconvolution approaches to mitigate the issue of marker co-expression across multiple cell types.

**Data Availability**

All data relevant to the study are included in the article or uploaded as supplementary information. The de-identified omics data generated in this study will be deposited in centralized public repositories as appropriate. Other data will be provided upon request from the corresponding author.

**Code Availability**

The open-source analysis software used in this study is publicly available and referenced as appropriate. No custom tools were developed in this study.

### Supplementary Figures and Figure Legends

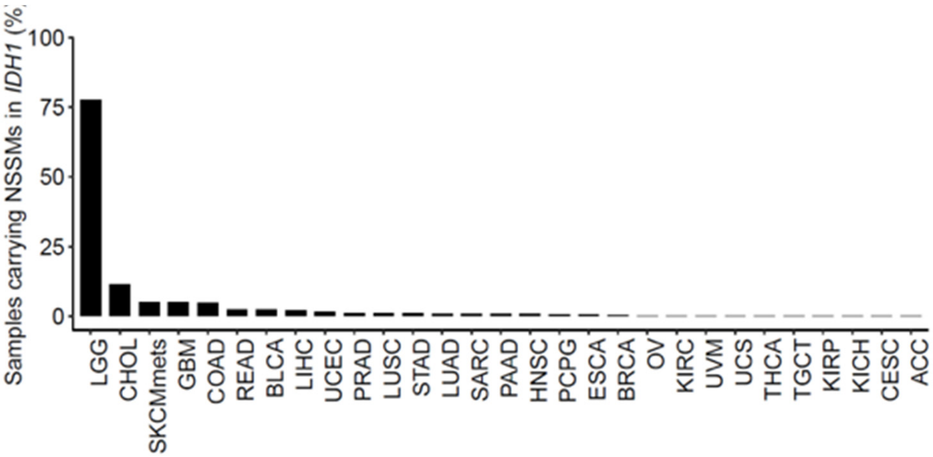


**Supplementary Figure 1. Distribution of IDH1 somatic mutations across human solid tumors from The Cancer Genome Atlas (TCGA).** *y*-axis shows the percentage of samples carrying non-synonymous somatic mutations (NSSM) in *IDH1* gene out of all tumor samples. *x*-axis denotes the TCGA study abbreviations for tumor types. ACC=Adrenocortical carcinoma; BLCA=Bladder Urothelial Carcinoma; LGG=Brain Lower Grade Glioma; BRCA=Breast invasive carcinoma; CESC=Cervical squamous cell carcinoma and endocervical adenocarcinoma; CHOL=Cholangiocarcinoma; COAD=Colon adenocarcinoma; ESCA=Esophageal carcinoma; GBM=Glioblastoma multiforme; HNSC=Head and Neck squamous cell carcinoma; KICH=Kidney Chromophobe; KIRC=Kidney renal clear cell carcinoma; KIRP=Kidney renal papillary cell carcinoma; LIHC=Liver hepatocellular carcinoma; LUAD=Lung adenocarcinoma; LUSC=Lung squamous cell carcinoma; OV=Ovarian serous cystadenocarcinoma; PAAD=Pancreatic adenocarcinoma; PCPG=Pheochromocytoma and Paraganglioma; PRAD=Prostate adenocarcinoma; READ=Rectum adenocarcinoma; SARC=Sarcoma; SKCMmets=Skin Cutaneous Melanoma (metastatic); STAD=Stomach adenocarcinoma; TGCT=Testicular Germ Cell Tumors; THCA=Thyroid carcinoma; UCS=Uterine Carcinosarcoma; UCEC=Uterine Corpus Endometrial Carcinoma; UVM=Uveal Melanoma.

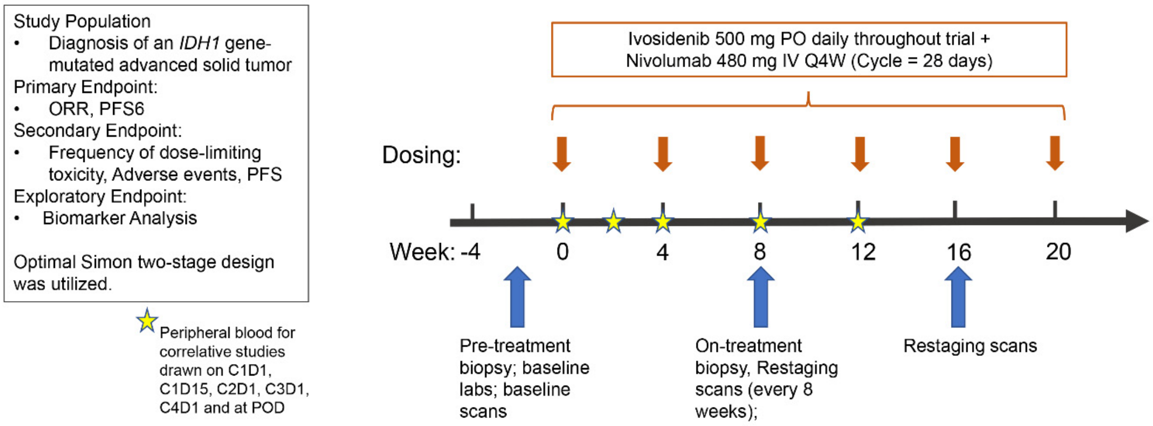


**Supplementary Figure 2. Study design schematic for the clinical trial of ivosidenib plus nivolumab in patients with IDH1 mutant advanced solid tumors.** Patients received daily oral ivosidenib (500 mg) and intravenous nivolumab (480 mg Q4W; 28-day cycles). Peripheral blood was collected for correlative studies at specified timepoints (C1D1, C1D15, C2D1, C3D1, C4D1, and progression). Tumor biopsies were obtained pre-treatment (Week 0) and on-treatment (Week 8), with imaging for response assessment performed every 8 weeks. The trial employed an optimal Simon two-stage design with primary endpoints of ORR and PFS6, secondary endpoints of toxicity and PFS, and exploratory biomarker analyses.

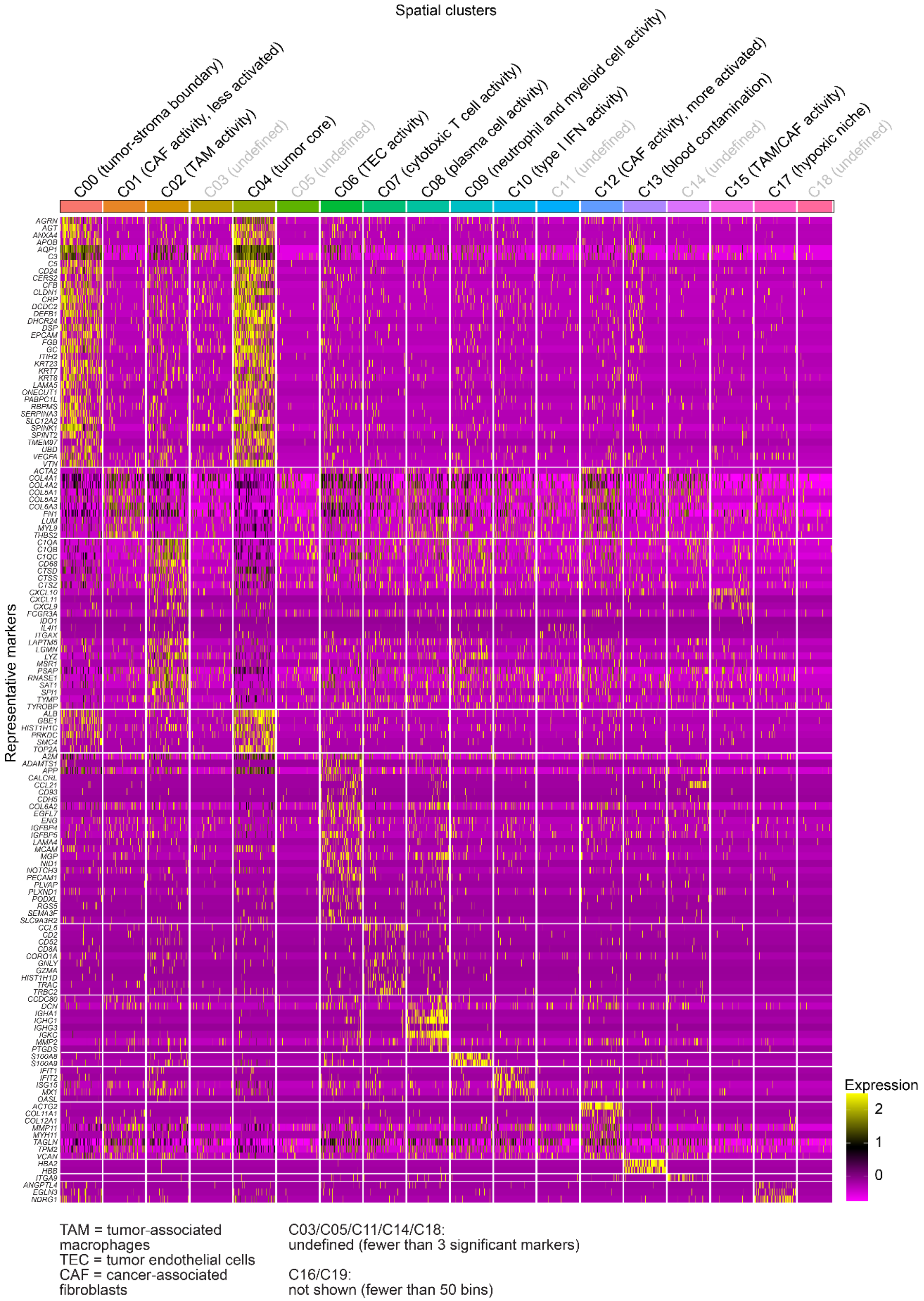


**Supplementary Figure 3. Spatial clusters from Visium HD spatial transcriptomics.**

Representative markers of each cluster are shown to the left of the heatmap. Biological annotation is shown next to each cluster label above the heatmap. 20 clusters were identified (C00-C19). Among those, 5 clusters (C03/05/11/14/18) were undefined with low numbers of significant markers; and 2 clusters (C16/19) were not shown due to low numbers of bins in each cluster. After excluding C13 where blood contamination was detected, 12 clusters were carried out for further analysis.

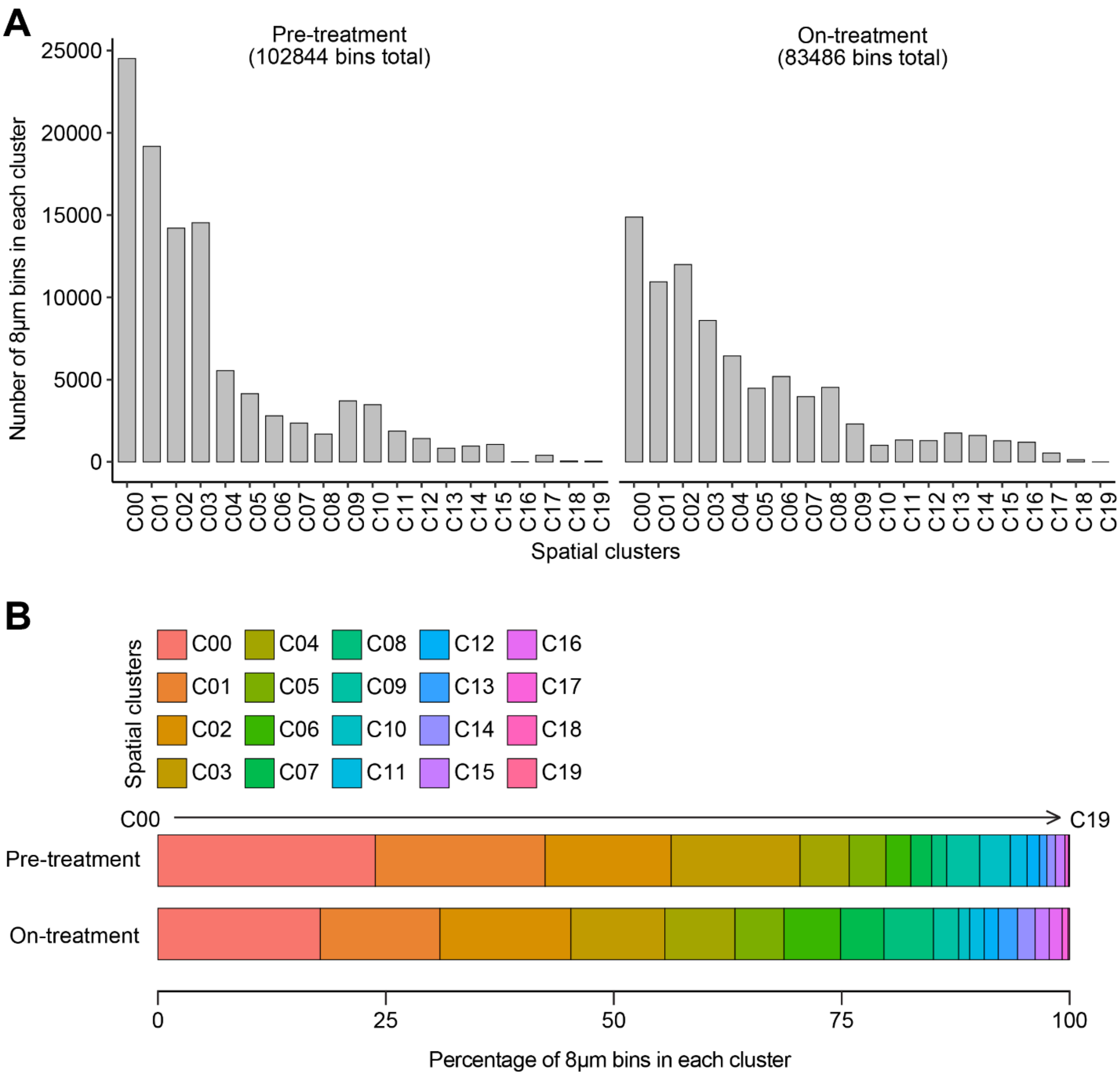


**Supplementary Figure 4. Distribution of bins across spatial clusters**. (**A**) Number of bins in each cluster from pre-treatment (left) or on-treatment tumor (right). (**B**) Percentage of bins in each cluster. 20 clusters (C00-C19) are shown.

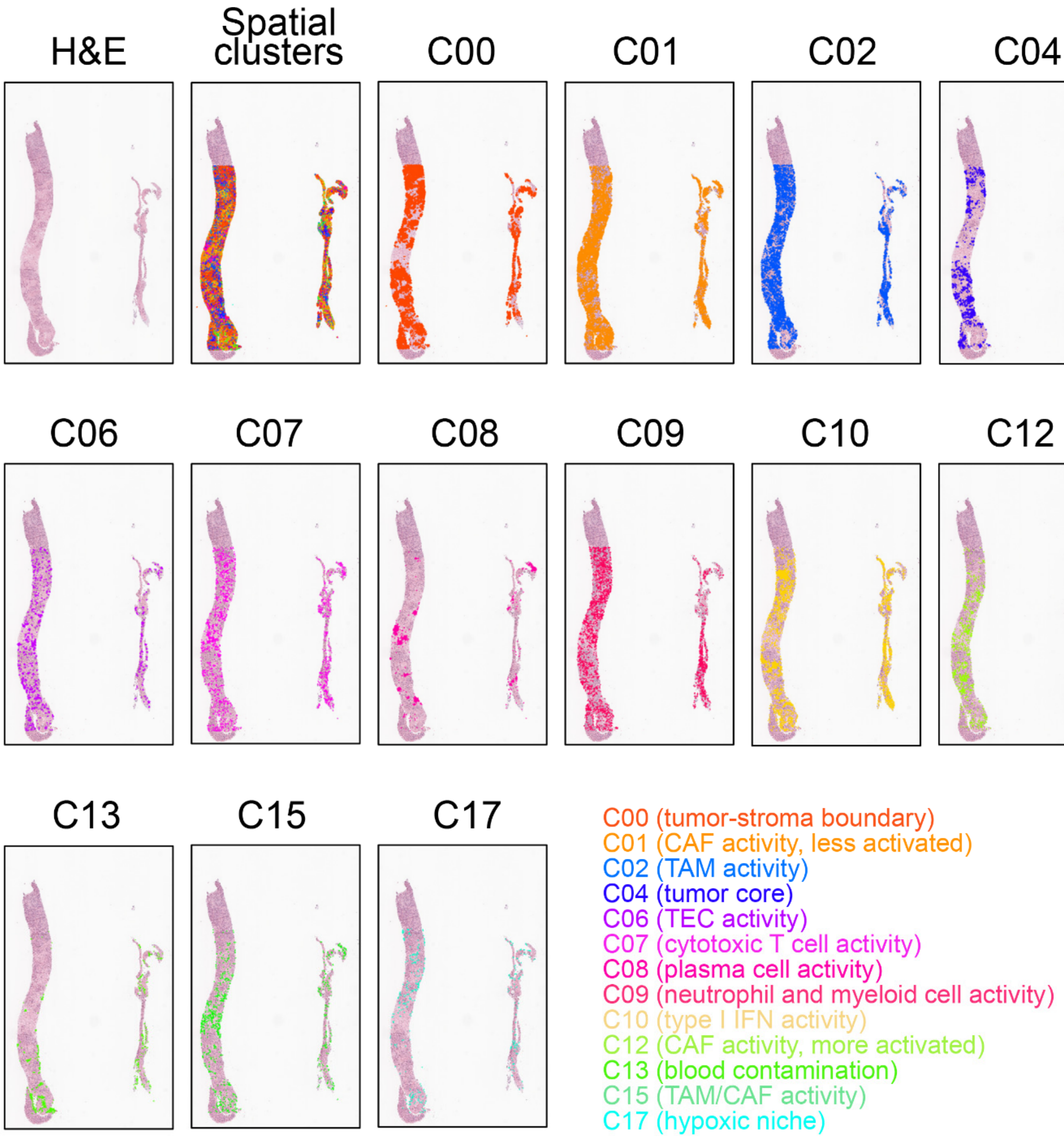


TAM = tumor-associated macrophages
TEC = tumor endothelial cells
CAF = cancer-associated fibroblasts

Clusters not shown:
C03/C05/C11/C14/C18 (undefined, fewer than 3 significant markers)
C16/C19 (fewer than 50 bins)

**Supplementary Figure 5. Spatial clusters in pre-treatment of patient 016**.

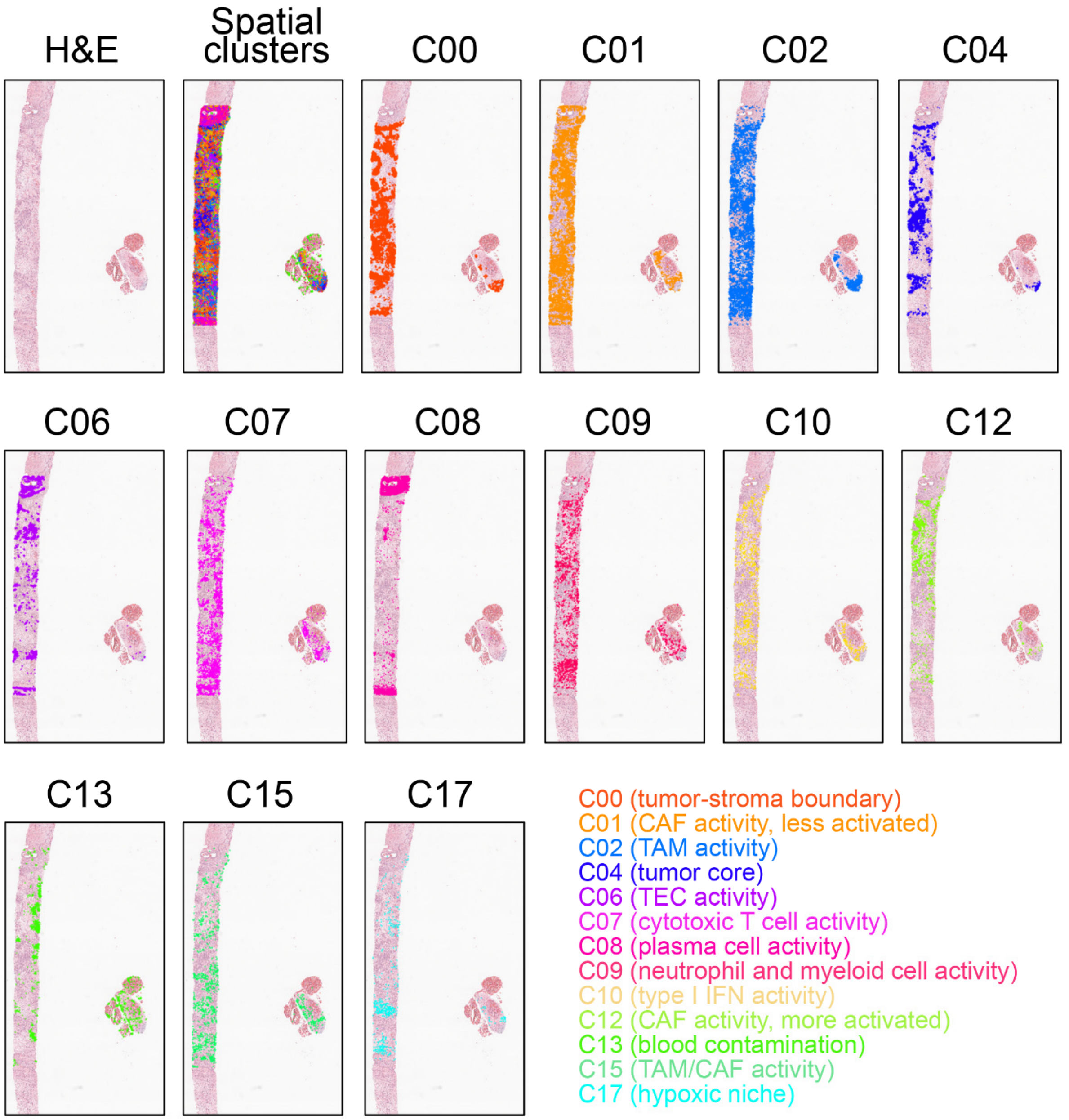


TAM = tumor-associated macrophages
TEC = tumor endothelial cells
CAF = cancer-associated fibroblasts

Clusters not shown:
C03/C05/C11/C14/C18 (undefined, fewer than 3 significant markers)
C16/C19 (fewer than 50 bins)

**Supplementary Figure 6. Spatial clusters in on-treatment of patient 016.**

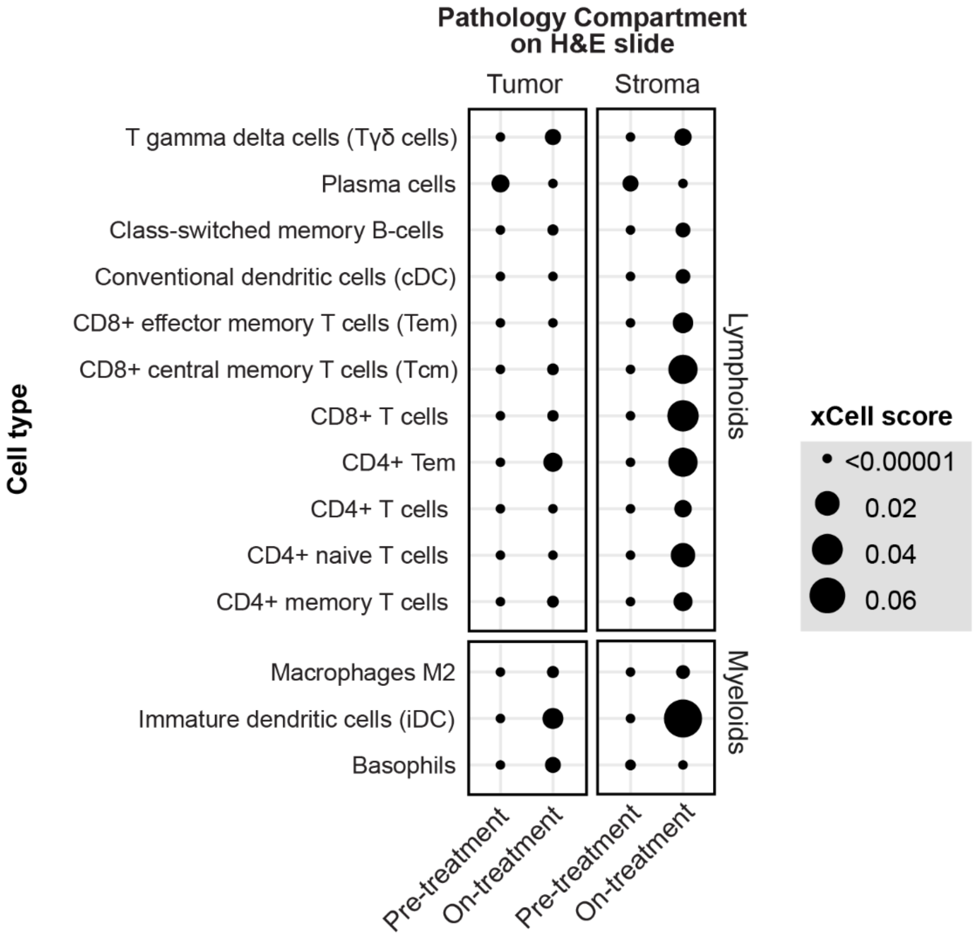


**Supplementary Figure 7. Digital cytometry of tumor and stroma compartment gene expression from pre- and on-treatment tumors by xCell**. Lymphoid and myeloid cell populations are shown to the left. Size of circle represents the enrichment scores from xCell analysis.

### Supplementary Tables

The titles are provided below; The tables are provided in separate spreadsheets.

**Supplementary Table 1. Summary of adverse events.**

**Supplementary Table 2. All adverse events.**

**Supplementary Table 3. Treatment-related adverse events.**
